# Association between increased anterior cingulate glutamate and psychotic-like symptoms, but not autistic traits

**DOI:** 10.1101/2023.02.01.23285183

**Authors:** Verena F Demler, Elisabeth F. Sterner, Martin Wilson, Claus Zimmer, Franziska Knolle

## Abstract

Despite many differences, autism spectrum disorder and schizophrenia spectrum disorder share environmental risk factors, genetic predispositions as well as neuronal abnormalities, and show similar cognitive deficits in working memory, perspective taking, or response inhibition. These alterations are already present in subclinical traits of these disorders. The literature proposes that alterations in the inhibitory GABAergic and the excitatory glutamatergic system could explain underlying neuronal commonalities and differences. Using magnetic resonance spectroscopy (^1^H-MRS), we investigated the associations between glutamate concentrations in the anterior cingulate cortex (ACC), the left/right putamen, and left/right dorsolateral prefrontal cortex and psychotic-like experiences (schizotypal personality questionnaire) and autistic traits (autism spectrum quotient) in 53 healthy individuals (28 women). To investigate the contributions of glutamate concentrations in different cortical and subcortical regions to symptom expression and their interactions, we used linear regression and moderation analyses. We found that glutamate concentration in the ACC but in none of the other regions predicted positive-like symptoms. None of the other clinical scores was associated with altered levels of glutamate. Specifying this finding, the moderation analysis showed that increased ACC glutamate was predictive of positive-like symptoms when glutamate concentrations in the right putamen were reduced, and that increased ACC glutamate was predictive of positive-like symptoms when disorganized traits were attenuated. This study provides evidence that an imbalance in the glutamatergic neurotransmitter system involving cortical and subcortical regions is linked to the expression of psychotic-like experiences, especially positive-like symptoms. These findings may facilitate the detection of individuals transitioning into an acute episode of psychosis.

## Introduction

Already in the earliest clinical reports of schizophrenia and autism spectrum disorder (ASD), their commonalities and associations have been discussed ^1–3^. On a phenomenological level, schizophrenia and ASD overlap regarding negative symptoms, social cognition, and perceptual alterations ^4^. Additionally, both disorders display similar cognitive deficits in terms of working memory function, perspective taking, or response inhibition ^5^. Nevertheless, ASD and schizophrenia show distinct clinical profiles and clinical progressions ^6, 7^. While ASD is a neurodevelopmental disorder with an early onset, usually diagnosed in childhood, and is characterized by a stable or improving long-term prognosis ^8^, schizophrenia typically develops later in adolescence or early adulthood and is associated with persistent long-term impairment ^9^. The greatest difference between the disorders, however, lies within the type of positive symptoms: While individuals with ASD may show repetitive or restricted behaviors, individuals with schizophrenia may suffer from hallucinations or delusions ^10^.

Despite those differences, both disorders share common environmental risk factors, genetic predispositions as well as neuronal abnormalities ^7, 11^. Patients with ASD and schizophrenia show, for example, abnormal development in the anterior cingulate cortex (ACC), striatum and frontal lobe ^12–14^. Additionally, shared atypical connections were identified in the default mode network and the salience network ^15^. Both disorders show similar alterations in reward processing ^16, 17^ and prediction error learning ^18–21^, with individuals with ASD showing greater impairment in social reward and prediction error learning. Importantly, both disorders show substantial overlap already at the subclinical and trait level ^22–24^.

Potential candidates providing an explanation for the underlying neuronal commonalities and differences are different neurotransmitter systems such as the glutamatergic system. The glutamate hypothesis of schizophrenia is well described ^25, 26^, building on the discovery that antagonists of a glutamate receptor (N-methyl-D-aspartate (NMDA) receptor) induced psychotic symptoms. Alterations in neurotransmitter systems may be assessed non-invasively with magnetic resonance spectroscopy (MRS). A recent review ^25^ summarizes that glutamate and Glx (glutamate+glutamine) concentrations are increased in the basal ganglia, thalamus and medial temporal lobe in schizophrenia patients. However, other studies reveal more inconsistent findings, showing increased prefrontal glutamate in anti-psychotic naïve patients ^27^, while another study ^28^ reports glutamate reductions in the ACC in psychosis, but could not detect alterations in the putamen or DLPFC. Reductions in glutamate concentrations have been found more consistently in the ACC in medicated early psychosis patients ^29^ and a mix of early psychosis and chronic schizophrenia patients ^30^. This supports the theory that increased glutamate is a sign of an acute psychotic or prodromal phase which builds up to a first episode and is linked to inflammatory processes ^31, 32^. Interestingly, there are strong links between altered levels of glutamate and neurofunction, which are task-dependent ^33^. In a recent meta-analysis, Zahid and colleagues ^34^ reported reduced positive associations between ACC glutamate levels and brain activity during resting state conditions, but increased positive associations between ACC glutamate levels and brain activity during cognitive control tasks in early psychosis patients. These studies seem to suggest that there are complex interactions, dependent on the brain state, between cortical and subcortical glutamate that may be altered in schizophrenia.

Also, in ASD research there is emerging evidence that an imbalance between different neurotransmitter systems, mainly the excitatory glutamatergic and inhibitory GABAergic system, contributes to the development of the disorder. Alterations with respect to glutamate have been reported in many cortical and subcortical regions. Horder and colleagues ^35^ for example found reduced levels of glutamate in the striatum to be associated with the severity of social symptoms in ASD. Interestingly, Page and colleagues ^36^ found increased concentrations of glutamate/glutamine in the amygdala/hippocampus, but not in parietal regions, while others found increased glutamate in the inferior frontal gyrus ^37^, the sensory motor cortex (children ^38^) or the putamen ^39^. Another study ^40^ found reduced glutamate concentrations in the auditory cortex and increased concentrations in the ACC. Alterations in these regions have also been reported by others ^41, 42^. However, also in ASD findings are inconsistent ^43–45^. Both disorders are associated with alterations in cortico-striatal-thalamic circuits ^46–48^ and abnormal excitatory glutamate neurotransmitter concentrations in overlapping cortical and subcortical areas ^28, 35, 49–53^ including the cingulate cortex, the prefrontal cortex and striatum. This suggests that a complex interaction of altered levels of glutamate in different cortical and subcortical regions may contribute to the development of symptoms within those two disorders.

Consistent with the continuum model of psychotic disorders, such as schizophrenia, and ASD, symptoms of schizophrenia and autism are reflected on a spectrum ranging from subclinical variants of psychotic-like experiences and autistic traits to clinical states of schizophrenia and ASD respectively ^54^ where schizophrenia and autism are seen as extremes of a continuum diverging in opposite directions from normality ^6, 55^. Frameworks of subclinical traits allow the investigation of neurobiological mechanisms underlying symptomatology without the influence of potential confounders such as medication, illness duration or age onset. Traits of these disorders may be assessed with the Schizotypal Personality Questionnaire (SPQ) and the Autism Spectrum Quotient (AQ) for psychotic-like experiences and autistic traits, respectively. As for the clinical stages of the disorders, strong associations between both subclinical spectra are reported in the literature. A recent meta-analysis ^56^ revealed a high correlation between autistic and global psychotic-like experiences of 0.48. Interestingly, however, the results indicate that while autistic traits share significant overlap with negative-like symptoms (*r*=0.54) and disorganized traits (*r*=0.36), they are only weakly associated with positive-like symptoms (*r*=0.26).

Only few studies explored changes of neurotransmitter concentrations associated to psychotic-like experiences and autistic traits. Ford and colleagues ^57^ investigated whether abnormalities in the glutamate/GABA ratio were linked to symptoms shared between the two traits and symptom severity using MRS. They found a positive correlation between glutamate/GABA ratio in the right superior temporal gyrus and the total scores of AQ, SPQ and AQ+SPQ. Additionally, positive correlations with subscales related to social skills and communication ^57^, indicating that alterations in glutamatergic and GABAergic neurotransmitter systems are associated with shared symptoms. Given that the literature reports alteration of neurotransmitter concentrations, mainly glutamate, across a number of different brain regions, it is likely that those regions contribute in an interactive way to the development of clinical symptoms.

The aim of the present study was, therefore, to use MRS to explore if and how glutamate concentrations in five cortical and subcortical areas interact and are associated with autistic traits and psychotic-like experiences in a sample of healthy individuals. The regions included the ACC, the left and right dorsolateral prefrontal cortex (DLPFC), and the left and right putamen. We hypothesized that increased levels of glutamate in the ACC and reduced levels of glutamate in the DLPFC and putamen would be associated with psychotic-like experiences, especially positive-like symptoms, and would not be moderated by autistic traits. On the other hand, we hypothesized that reduced levels of ACC glutamate and increased levels of glutamate in the putamen would be associated with autistic traits which would be, at least, partially mediated by psychotic-like experiences. Potentially moderated interactions of subcortical and cortical glutamate concentrations on subclinical scores were tested with moderation analyses.

## Patients and methods

### Participants and subclinical questionnaires

53 healthy subjects (28 women) aged 18-35 years participated in this study. See supplementary materials for recruitment and inclusion criteria. The study was approved by the medical research ethics committee of the Technical University of Munich. All subjects gave written informed consent in accordance with the Declaration of Helsinki.

All participants completed a German version ^58^ of the Schizotypal Personality Questionnaire (SPQ) ^59^ capturing psychotic-like experiences which may be factored into positive-like symptoms, negative-like symptoms, and disorganized traits ^60^; as well as the Autism Spectrum Quotient (AQ) ^61^ assessing autistic traits (Table 1, Figure S1; see supplementary material for details).

**Table 1:** Demographic data and clinical scores.

|  | Female (n = 28) | Male (n = 27) | P-value<sup>1</sup> | W |
| --- | --- | --- | --- | --- |
| Age | 23.54 (3.53) | 23.93 (4.21) | 0.9392 | 373 |
| SPQ: positive-like symptoms (/132) | 34.04 (23.80) | 28.70 (25.20) | 0.3080 | 439 |
| SPQ: negative-like symptoms (/100) | 36.89 (15.65) | 34.33 (20.49) | 0.4535 | 423 |
| SPQ: disorganized traits (/64) | 21.93 (14.57) | 21.22 (13.12) | 0.9933 | 379 |
| AQ: total score (/50) | 21.21 (7.03) | 20.96 (7.65) | 0.8794 | 387.5 |

*Note: Values are mean (SD)*
*SPQ, Schizotypal Personality Questionnaire; AQ, Autism Spectrum Quotient*
<sup>1</sup>Wilcoxon rank sum test

### MR Image acquisition

Structural MRI and ^1^H-MRS data were collected on a 3T Philips Ingenia Elition X MR-Scanner (Philips Healthcare, Best, The Netherlands) using a 32-channel head coil. For anatomical reference and spectroscopic voxel placement, we acquired a T1-weighted magnetization prepared rapid gradient echo (MPRAGE) sequence: echo time (TE)=4ms, repetition time (TR)=9ms, Flip angle (α)=8°, shot interval=3000ms, slice number=170, matrix size=240×252 and voxel size=1×1×1mm³. MRS data were collected using a ¹H-MRS single voxel ECHO volume Point Resolved Spectroscopy Sequence (PRESS) sequence: TR = 2000ms, TE set to shortest (35.6ms-41.2ms), samples = 1024, bandwidth = 2000Hz, phase cycle steps = 16, and flip angle = 90°. We used the conventional Philips water suppression technique (excitation) that performs Automatic Water Suppression Optimization (AWSO) prescans to minimize the residual water (window=140Hz, second pulse angle=300). See supplementary materials for further descriptions. We placed one voxel in the ACC (20×20×20mm) and one voxel per hemisphere in the Putamen (20×15×20mm) as well as in the DLPFC (30×20×20mm); see Figure 1A for voxel placement overlap.

**Figure 1:**
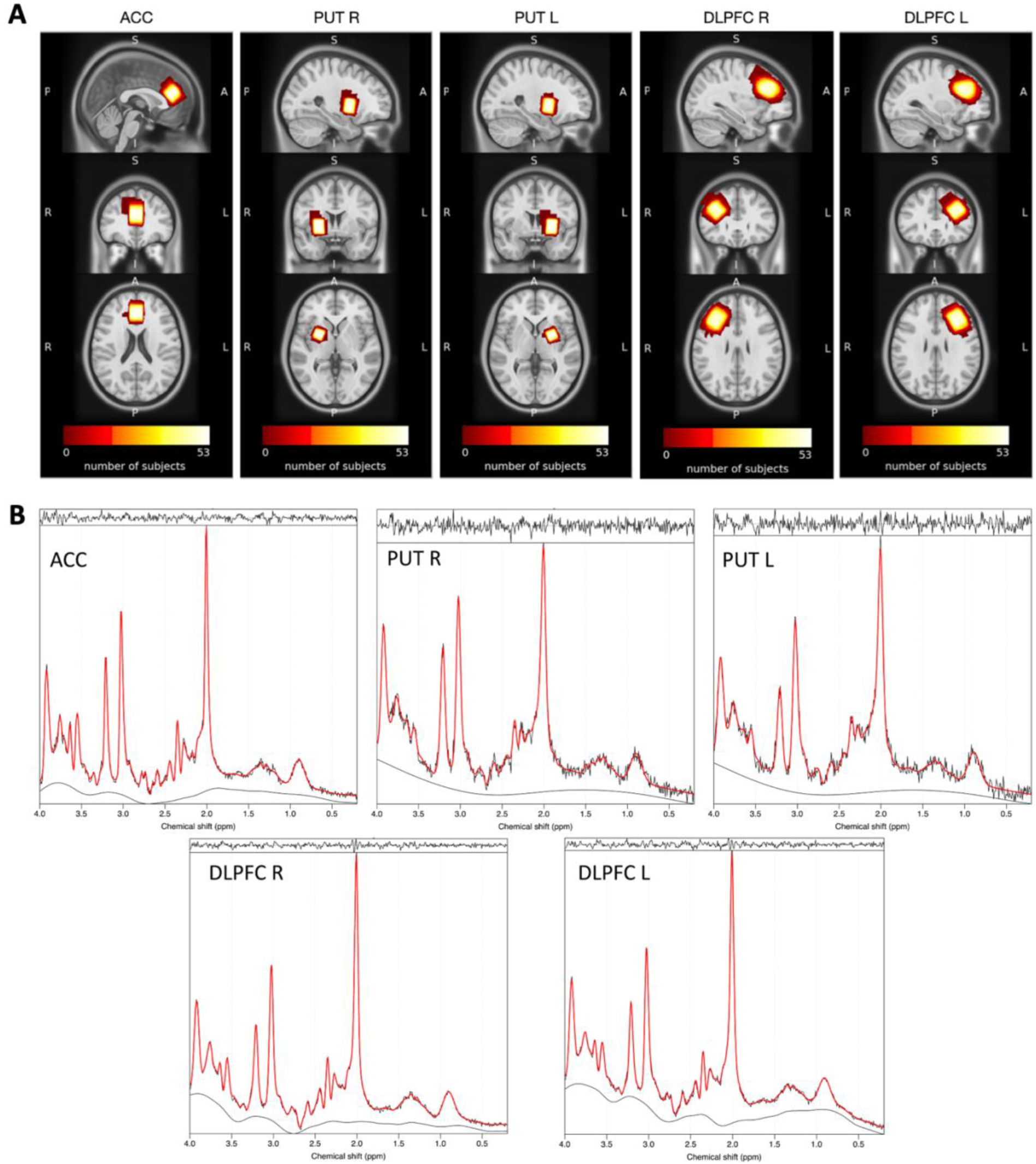
Voxel placement and representative fitted MRS spectra. Note: (A) Placement of MRS voxels in the ACC, putamen, and DLPFC. The colors indicate the areas covered by the subjects’ individually placed MRS voxels. The individual voxels were standardized with SPM, overlapped in MRIcroGL and visualized in FSLeyes. (B) ^1^H-MRS spectrum fitted by spant. ACC, anterior cingulate cortex; PUT R, right putamen; PUT L, left putamen; DLPFC R, right dorsolateral prefrontal cortex; DLPFC L, left dorsolateral prefrontal cortex

### ^1^H-MRS Analysis

The MRS data were analyzed using Spectroscopy Analysis Tools (spant) version 2.6.9 ^62^ (https://martin3141.github.io/spant/index.html), an open-source R toolbox. We aligned the spectrum to the total N-acetylaspartate (NAA) resonance at 2.01 ppm and removed the residual water signal using an HSVD filter for each participant spectrum. Beforehand, the automatic processing of the scanner had already performed coil combination and phase-frequency alignment, before averaging each repetition.

As the TE of our MRS data varied, we simulated six different PRESS basis sets for each TE (36, 37, 38, 39, 40, 41) with a bandwidth of 2000Hz and 1024pts using MARSS in INSPECTOR version 11-2021 ^63^. Based on recommendations in the LCModel manual ^64^, we selected the following metabolites: alanine, aspartate, creatine (Cr), GABA, glucose, glutamate (Glu), glutamine, glutathione, glycerophosphocholine, lactate, myoinositol, NAA, N-acetyl-aspartylglutamate (NAAG), phosphocholine, phosphocreatine (PCr), scyllo-inositol and taurine. To this basis set we added the default macromolecular and lipid components provided by spant.

The fitting in spant uses an adaptive baseline fitting algorithm (ABfit) ^65^, which accurately estimates the optimal baseline. For the quantification of absolute metabolite concentrations, given in mMol/kg tissue water, calculated tissue volume fractions within the MRS voxel are needed. Therefore, we used svs_segment from fls_mrs version 2.0.2 ^66^ (https://open.win.ox.ac.uk/pages/fsl/fsl_mrs/), which relies on fsl_anat to run FSL FAST tissue segmentation. The structural T1 image was segmented into gray matter (GM), white matter (WM), and cerebrospinal fluid (CSF). The segmentation parameters are saved in a JSON-file. We then ran the spant::scale_amp_molal_pvc() with the ABfit method to quantify glutamate, whereas correction for tissue fractions is based on Gasparovic and colleagues ^67^. See supplementary materials for a detailed workflow diagram (Figure S2). The fit of the glutamate spectra was appropriate for all participants (Figure 1B). Spectral exclusion criteria were either visual failure of the fitting algorithm or Cramer-Rao lower bounds (CRLB) > 20% of glutamate concentration. No data had to be excluded.

### The association between autistic traits and schizotypal traits

To investigate the relationship between the SPQ subscales and autistic traits, we calculated Spearman’s rank correlation coefficients as the assumptions of normality, linearity and homoscedasticity were not met. The correlation analyses were performed in R using the Hmisc package version 4.7.1 (https://hbiostat.org/R/Hmisc/).

### Linear and moderated association between Glutamate and clinical scores

To understand the contributions of glutamate concentrations in different cortical and subcortical regions to symptom expression, we first conducted four linear regression models with symptom score (i.e., positive-like symptoms, negative-like symptoms, disorganized traits, autistic traits) as outcome and all five glutamate concentrations as predictor variables We also conducted the linear regression model with total Cr (tCr; Cr + PCr) and total NAA (tNAA; NAA + NAAG) to test whether the results were specific to glutamate alterations. These two metabolites were chosen because they have singlet resonances as well as high concentrations, and therefore a smaller error estimate for quantification.^68^ We then used two sets of moderation analyses to investigate, first, whether the association between the significantly predicted clinical score and the significant glutamate concentration was moderated by the other glutamate concentrations, and second, whether the association between the significantly predicted clinical score and the significant glutamate concentration were moderated by the other clinical scores. The significance level was set to p=0.05. Bonferroni multiple comparison corrections were applied, corrected significance levels are reported together with the results. Analyses were performed in R using the stats package version 4.0 (https://stat.ethz.ch/R-manual/R-devel/library/stats/html/00Index.html).

## Results

### ^1^H-MRS Glutamate levels and spectral quality

We measured the concentration of glutamate in five voxels of interest, the ACC, the left putamen, right putamen, left DLPFC and right DLPFC. For all subjects, the glutamate concentration could be estimated. All acquired spectra were of good quality. Results are presented in Table 2.

**Table 2:**
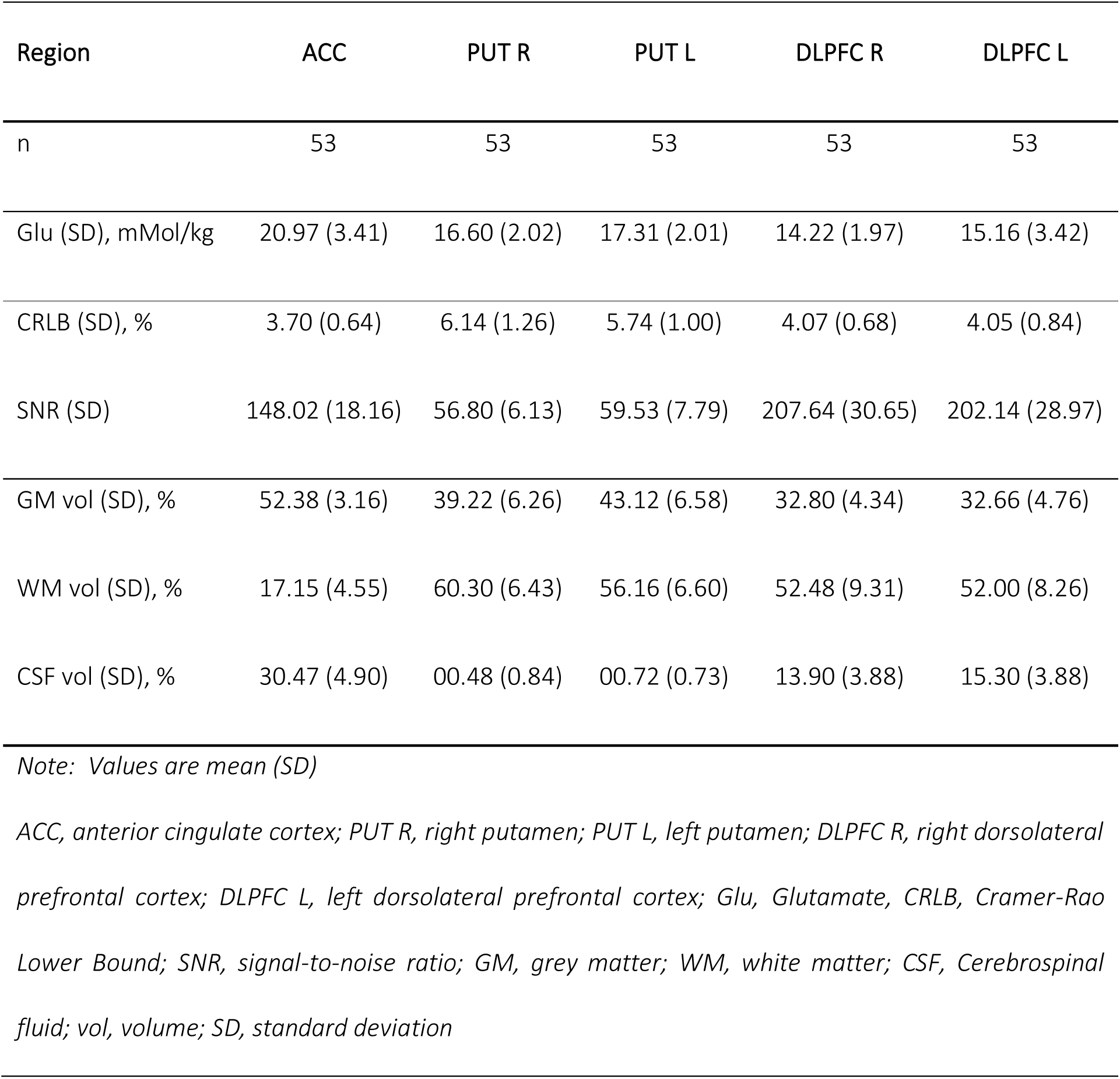
¹H-MRS quality parameters and glutamate levels by region.

### Correlation between the subscores of SPQ and AQ

Using the Spearman’s rank correlation coefficient, we found a strong, significant positive correlation between positive-like symptoms and disorganized traits (r=0.76, p<0.00001) and between negative-like symptoms and autistic traits (r=0.73, p<0.00001), a moderate, significant correlation between negative-like symptoms and the disorganized traits (r=0.64, p<0.00001), as well as between positive-like symptoms and the negative-like symptoms (r=0.45, p<0.0001). Lastly, we observed a weak, significant correlation between disorganized traits and autistic traits (r=0.39, p=0.0039) and a weak correlation between autistic traits and positive-like symptoms (r=0.19, p=0.17). Correlation strength was classified according to Akoglu ^69^. All significant correlations survived Bonferroni multiple comparisons correction (corrected p-value=0.0083). The correlations between the subscores are shown in Figure 2. The chord diagram was created in R using the circlize-package Version 0.4.15 ^70^.

**Figure 2:**
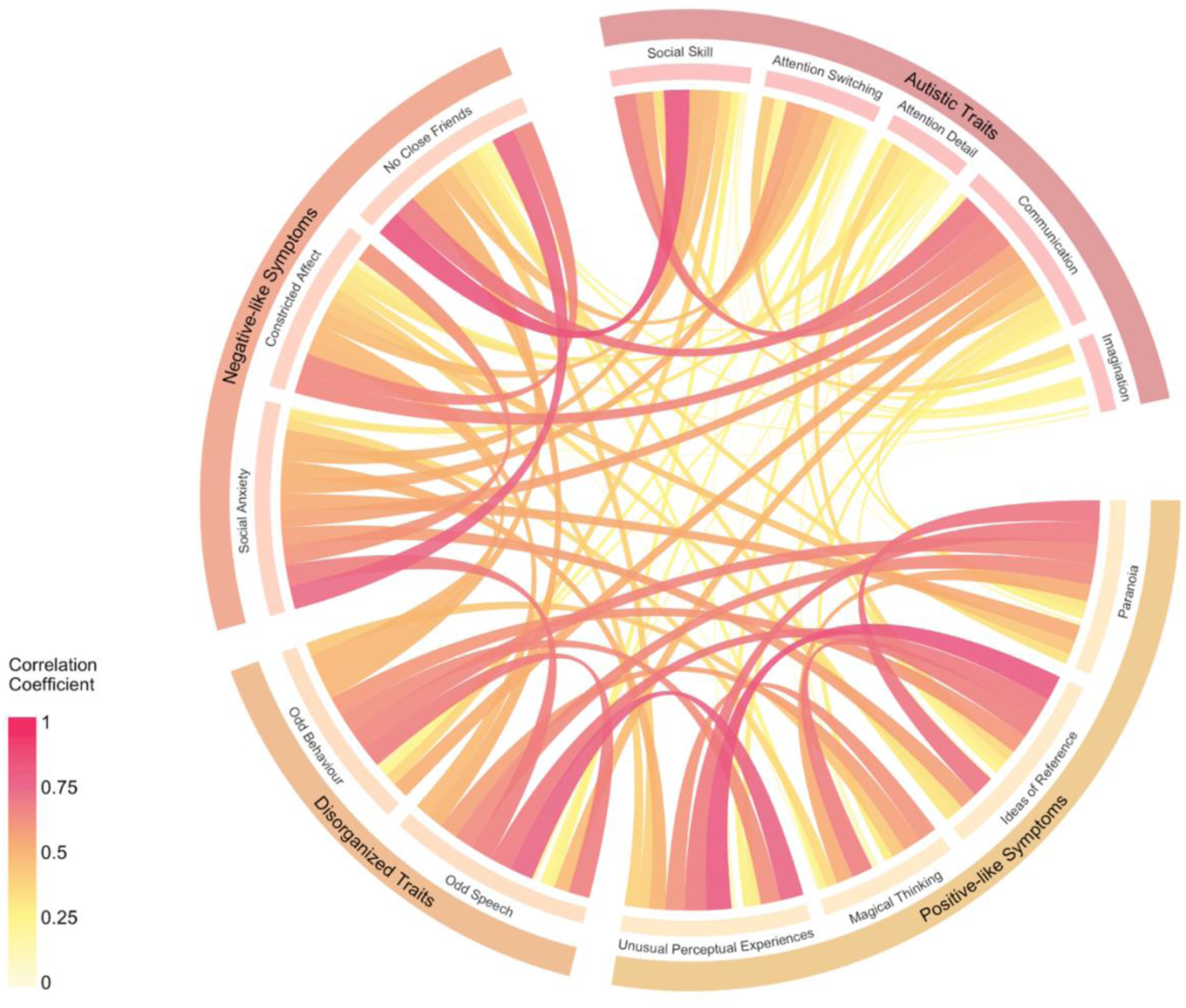
Visualization of the correlations between the subscores Note: The Chord Diagram describes the relationship between the individual items of the subscores of the AQ and SPQ using Spearman Correlation Coefficients. The subscores are structured according to their respective associated traits (i.e., Autistic Traits for AQ; Negative-like Symptoms, Posititive-like Symptoms, and Disorganized traits for SPQ). The color and width of the bidirectional links indicate the strength of the connection between each of the subscores – the wider and darker the stronger.

### Association between positive-like symptoms and glutamate concentration in the anterior cingulate cortex

We first fitted a complete multiple linear regression model to test if glutamate concentrations in our five voxels of interest predicted symptom scores. We found that positive-like symptoms (F(47,5)=1.55, p=0.192, r^2^=0.05) were significantly predicted by glutamate concentration in the ACC (β=2.25(0.98), t=2.30, p=0.026). These results were specific to glutamate alterations, as shown in control analyses using tNAA and tCr (Table S2). There was no association between negative-like symptoms, disorganized traits, or autistic traits and cortical or subcortical glutamate concentration. In a further control analysis, we confirmed that alterations in ACC glutamate were independent of changes in grey matter volumes in the respective region (Figure S3).

We then computed two sets of moderation analyses. In the first set, we investigated possible moderation effects of the glutamate concentrations in the left and right DLPFC and putamen on the significant interaction between ACC glutamate concentration and positive-like symptoms. We found a significant interaction effect between the glutamate concentration in ACC and the right putamen on positive-like symptoms (F(49,3)=4.97, p=0.004, r^2^=0.19, ACC: β=28.94(9.21), t=3.14, p=0.003, right putamen: β=32.09(10.94), t=2.93, p=0.005, ACC*right putamen: β=-1.59(0.55), t=-2.91, p=0.006), revealing a partial moderation of glutamate concentrations in the right putamen on the interaction of ACC glutamate and positive-like symptoms. This moderation effect indicates that high levels of ACC glutamate are predictive of psychotic symptoms when glutamate concentrations in the right putamen are decreased (Figure 3A). The effect survived Bonferroni multiple comparison correction (level of significance, p<0.0125). None of the other glutamate concentrations showed a significant moderation effect.

**Figure 3:**
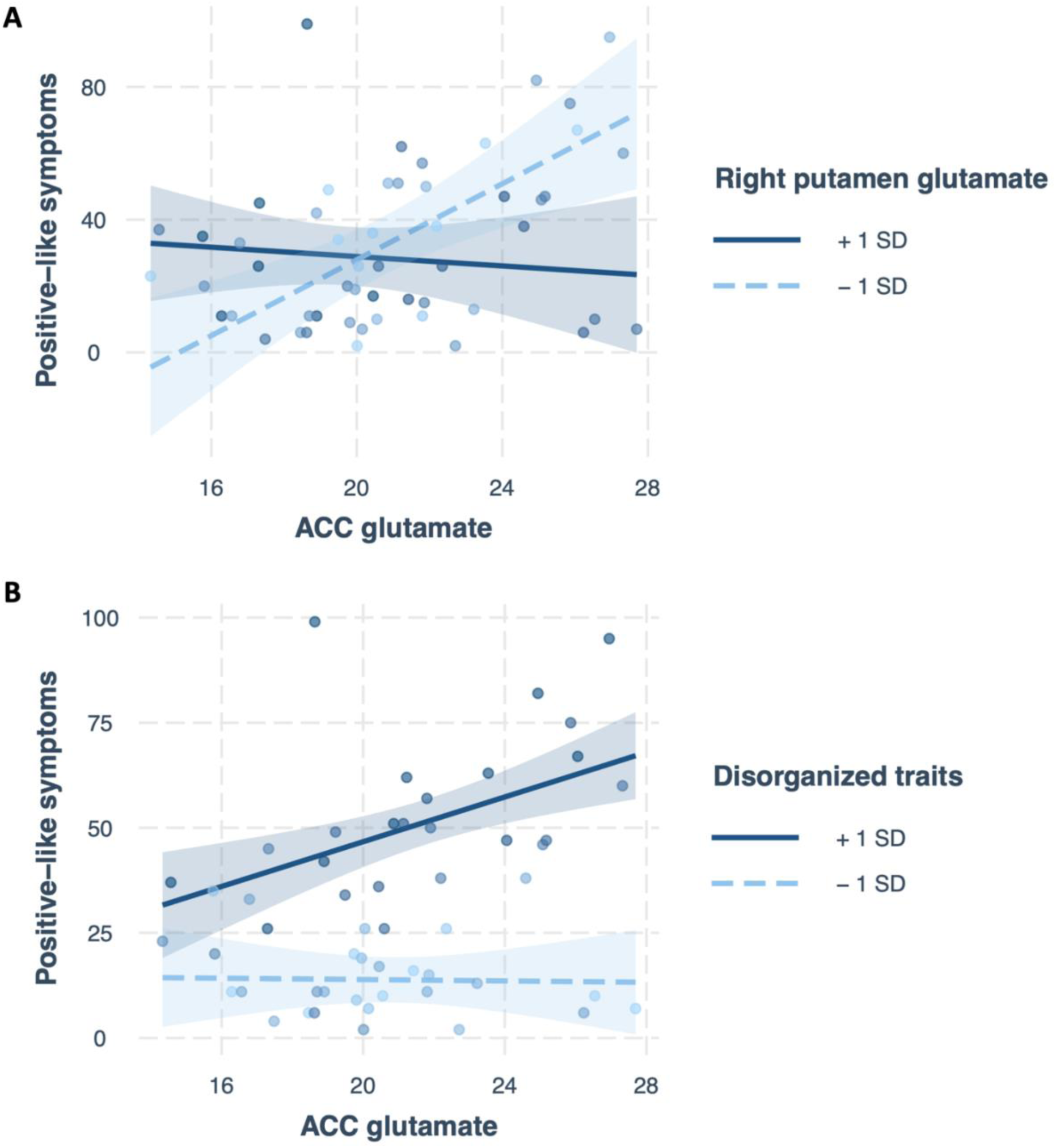
Moderation effects on the interaction of ACC glutamate and positive-like symptoms Note: (A) Significant moderating effect of decreased concentrations of glutamate in the right putamen (-1SD, dashed light blue line) on the interaction between ACC glutamate and positive like symptoms. (B) Significant moderating effect of increased disorganized traits (+1SD, solid dark blue line) on the interaction between ACC glutamate and positive like symptoms.

In the second set of moderation analyses, we investigated possible moderation effects of the different symptom traits, negative-like symptoms, disorganized traits and autistic traits, on the significant interaction between ACC glutamate concentration and positive-like symptoms. We found a significant interaction effect between the glutamate concentration in ACC and disorganized traits on positive-like symptoms (F(49,3)=39.23, p<0.001, r^2^=0.69, ACC: β=-0.95 (1.08), t=-0.88, p=0.33, disorganized traits: β=-0.81(0.87), t=-0.93, p=0.38, ACC*disorganized traits: β=0.10(0.04), t=2.54, p=0.014), revealing a moderation of disorganized traits on the interaction of ACC glutamate and positive-like symptoms. This moderation effect indicates that high levels of ACC glutamate are predictive of psychotic symptoms when disorganized traits are increased (Figure 3B). The effect survived Bonferroni multiple comparison correction (level of significance, p<0.017). None of the other symptom subscores showed a significant moderation effect.

## Discussion

In this study, we explored if and how glutamate concentrations in five cortical and subcortical areas interact and are associated with autistic traits and psychotic-like experiences in a sample of healthy individuals. We found that glutamate concentrations in the ACC, but in none of the other regions, predicted positive-like symptoms in healthy individuals. None of the other clinical scores was associated with altered levels of glutamate. Using two sets of moderation analyses, we furthermore found that, on the one hand, high levels of ACC glutamate are predictive of psychotic symptoms when glutamate concentrations in the right putamen were reduced, and that, on the other hand, high levels of ACC glutamate are predictive of psychotic symptoms when disorganized traits were increased.

### Association between ACC glutamate and positive-like symptoms

ACC glutamate alterations have been associated with psychotic-like experiences in healthy individuals ^71^, symptoms and structural changes in high-risk individuals ^72^, symptoms in first episode psychosis ^72–74^ and chronic psychosis ^75^. However, findings are inconsistent with some finding higher levels ^31, 73^, while others report reductions ^29, 30^ or no differences ^76, 77^. Although Modinos and colleagues ^77^ did not find different levels of glutamate in the ACC between individuals with high schizotypy and low schizotypy, they found that increased grey matter volume in ACC negatively related to ACC glutamate, supporting the view that glutamatergic alteration may explain structural changes which are associated with the development of psychotic-like experiences. The association in this one, but also other studies ^71^, however, suggests that decreasing levels of glutamate in the ACC are associated with specific pathologies in psychosis, whereas our study indicates that higher levels of ACC glutamate are associated with development of psychotic-like experiences. Our results correspond to findings from Demro and colleagues ^78^ who reported that increased subclinical symptoms of grandiosity were linked to increased levels of glutamate in the ACC. Egerton and colleagues ^79^ found higher levels of glutamate/creatine-ratio in the anterior cingulate cortex in symptomatic first episode psychosis patients compared to those in remission. In a different study, Egerton and colleagues ^80^ found that higher levels of glutamate metabolites in the ACC predicted treatment response, indicating that higher baseline levels are associated with poorer treatment response, while lower levels of ACC glutamate are predictive of improvements across positive and negative symptoms as well as general functioning in first episode psychosis patients. Relatedly, Godlewska and colleagues ^28^ reported lower levels of glutamate in the ACC in early psychosis patients compared to healthy controls. Importantly, however, the majority of patients included in this study were medicated, which confirms results from a recent meta-analysis ^81^ on the impact of anti-psychotic treatment on frontal glutamate levels. The authors found that antipsychotics were linked to a significant decrease in frontal Glx levels in both first episode and chronic schizophrenia patients, with the effect being stronger in first episode psychosis patients. As our results indicate that ACC glutamate levels are specifically predictive of positive-like symptoms but not of any other subclinical traits, the results are in line with the theory that increased glutamate is a sign of an acute psychotic or prodromal phase which builds up to a first episode and is linked to inflammatory processes ^31, 32^. The theory suggests that a hypofunction of NMDA receptors reduces the activity of inhibitory GABAergic inter-neurons, on which they are located. This activity reduction increases glutamatergic neurotransmission of pyramidal cells, and may contribute to the development of, especially, positive symptoms during an acute episode of psychosis_32,82_.

### Moderation effect of reduced glutamate in right putamen on association between ACC glutamate and positive-like symptoms

Although, glutamate concentrations in the other regions did not predict trait symptom expressions, we are able to show that glutamate levels in the right putamen significantly moderated the association between ACC glutamate and positive-like symptoms. Notably, our results are unconfounded by medication or disease duration, possibly indicating that it is not only an imbalance between excitatory and inhibitory processes that are contributing to the development of psychotic-like symptoms ^50, 83^, but also the interaction of different cortical and subcortical regions ^47, 48^. Interestingly, there are a number of studies showing altered connectivity between the putamen and the ACC in different stages of the disease ^50, 84^. Shukla and colleagues ^50^ found that the functional connectivity pattern of the ACC was correlated with glutamate, but also GABA, concentrations. Similar results have been reported by Overbeek and colleagues ^83^, who showed that aberrant associations of ACC glutamate and of ACC GABA with ACC functional connectivity was significantly different in first-episode psychosis patients compared to controls in several cortical and subcortical regions including the putamen. Although this study provides clearer evidence for a modulatory effect of GABA on the connectivity between ACC and striatal regions, it generally indicates the impact of the glutamatergic and GABAergic neurotransmitter system on functional connectivity. Moreover, Modinos and colleagues ^71^ showed that in individuals with high schizotypy (i.e., increased psychotic-like experiences) ACC glutamate correlated significantly and negatively with striatal activation including the putamen, providing further evidence for the modulatory effect on functional and structural alterations possibly contributing the development of symptoms. Nevertheless, we would like to point out that the current results are inconsistent most likely due to the use of various individuals at various stages of the disease including pre-clinical stages.

### Moderation effect of increased disorganized traits on association between ACC glutamate and positive-like symptoms

In addition to the moderation effect of glutamate in the right putamen, we found that the association between ACC glutamate and positive-like symptoms was strengthened when disorganised traits were increased in the individuals. Considering the strong positive correlation between positive-like symptoms and disorganized traits in general in individuals with psychotic-like experiences ^56^, but especially in our sample (r=0.76, p<0.00001), is seems little surprising. This result however may further support the hypothesis that increased levels of ACC glutamate are indicative of an acute phase, as both subclinical scores have been found to be associated with positive symptoms in schizophrenia spectrum disorder ^85^, which are also more prominent in an acute phase ^25^.

### Lack of effect for other regions and subclinical traits

Surprisingly, our data did not reveal any associations between different glutamate concentrations and other subclinical symptoms than positive-like symptoms. Generally, the literatures present an inconsistent picture regarding glutamate concentration in different regions. For example, although Godlewska and colleagues ^28^ reported alterations in glutamate in the ACC in early psychosis patients compared to healthy controls, they did not find differences in the putamen and DLPFC. However, especially the literature with regard to subclinical schizotypal traits is sparse, and mainly concentrates on ACC and hippocampal glutamate ^71, 77^. To our knowledge only one other study investigated prefrontal glutamate in individuals with high schizotypy compared to low schizotypy ^86^, in which the authors reported a reduction in high schizotypy.

The literature is similarly sparse with regard to associations between glutamate and subclinical autistic traits. Data by Ford and colleagues ^87^ suggest social disorganisation is associated with increased glutamate/GABA+ ratio in the right superior temporal region across both the autistic and schizotypal spectrum. In contrast, Kondo and colleagues ^88^ report that autistic and schizotypal traits were associated with the Glutamine+Glutamate/GABA ratio in the auditory cortex but not in the frontal areas, including ACC, DLPFC and inferior frontal cortex. More research is needed, especially focusing on consistent ways of measuring metabolite differences, as it is not only the choice of location for voxel placement that differs grossly among studies but also the choice of metabolite representation (e.g., glutamate/GABA ratio, absolute glutamate, glutamate/creatine ratio, etc) and selection of basis sets.

### Limitations

This study has several limitations. First, we did not analyze levels of GABA in the voxels of interest, as no spatial editing was applied to the MRS sequence. Future studies should try to measure both metabolites reliably in order to understand interactions between neurotransmitters and regions. Second, we were unable to recruit participants with highly increased scores along both spectra. One possible explanation is that many individuals with highly increased scores would already have a clinical psychiatric diagnosis, which was an exclusion criterium in our study. Nevertheless, follow-up studies should recruit larger samples allowing for a larger spectrum. Third, our sample does not include many individuals with low SPQ and high AQ scores, and vice versa, which would be necessary in order to clearly differentiate between clinical traits. Yet, our sample represents the distribution of subclinical symptoms within the general population as confirmed by similar correlations of subclinical subscores compared to previous studies ^56, 89^.

## Conclusion

Taken together, this study shows that an interaction between glutamate in the ACC and the putamen is specifically associated with positive-like symptoms, which indicates that an imbalance in the glutamatergic neurotransmitter system involving cortical and subcortical regions may contribute to the development of psychotic-like experiences, especially positive-like symptoms. These findings may ultimately facilitate early detection of individuals transitioning into an acute episode of psychosis.

## Data Availability

Data is available upon reasonable request to the authors.

## Acknowledgments

We would like to thank all participants for their time and engagement.

This work was supported by the doctoral program “Translationale Medizin” of the Technical

University of Munich funded by Else Kröner-Fresenius-Stiftung (EKFS) to VD.

## Conflict of interest

The authors declare no conflict of interest.

**Ethics:** The medical research ethics committee of the Technical University of Munich gave ethical approval for this work. All subjects gave written informed consent in accordance with the Declaration of Helsinki.

## References

1 Mercier CA. Sanity and insanity. Scott: London, 1890.

2 de Lacy N, King BH. Revisiting the Relationship Between Autism and Schizophrenia: Toward an Integrated Neurobiology. Annu Rev Clin Psychol 2013; 9: 555–587.

3 Bleuler E. Dementia praecox, oder, Gruppe der Schizophrenien. Wellcome Collect. 1911.https://wellcomecollection.org/works/v2qqa9g4/items (accessed 20 Jan2023).

4 Jutla A, Foss-Feig J, Veenstra-VanderWeele J. Autism spectrum disorder and schizophrenia: An updated conceptual review. Autism Res 2022; 15: 384–412.

5 Ford TC, Apputhurai P, Meyer D, Crewther DP. Cluster analysis reveals subclinical subgroups with shared autistic and schizotypal traits. Psychiatry Res 2018; 265: 111–117.

6 Nenadić I, Meller T, Evermann U, Schmitt S, Pfarr J-K, Abu-Akel A et al. Subclinical schizotypal vs. autistic traits show overlapping and diametrically opposed facets in a non-clinical population. Schizophr Res 2021; 231: 32–41.

7 Zheng Z, Zheng P, Zou X. Association between schizophrenia and autism spectrum disorder: A systematic review and meta-analysis. Autism Res 2018; 11: 1110–1119.

8 Anderson DK, Liang JW, Lord C. Predicting young adult outcome among more and less cognitively able individuals with autism spectrum disorders. J Child Psychol Psychiatry 2014; 55: 485–494.

9 McCutcheon RA, Reis Marques T, Howes OD. Schizophrenia—An Overview. JAMA Psychiatry 2020; 77: 201.

10 Trevisan DA, Foss-Feig JH, Naples AJ, Srihari V, Anticevic A, McPartland JC. Autism Spectrum Disorder and Schizophrenia Are Better Differentiated by Positive Symptoms Than Negative Symptoms. Front Psychiatry 2020; 11.

11 Jutla A, Foss-Feig J, Veenstra-VanderWeele J. Autism spectrum disorder and schizophrenia: An updated conceptual review. Autism Res 2022; 15: 384–412.

12 Langen M, Schnack HG, Nederveen H, Bos D, Lahuis BE, de Jonge MV et al. Changes in the Developmental Trajectories of Striatum in Autism. Biol Psychiatry 2009; 66: 327–333.

13 McCutcheon RA, Abi-Dargham A, Howes OD. Schizophrenia, Dopamine and the Striatum: From Biology to Symptoms. Trends Neurosci 2019; 42: 205–220.

14 Cauda F, Costa T, Nani A, Fava L, Palermo S, Bianco F et al. Are schizophrenia, autistic, and obsessive spectrum disorders dissociable on the basis of neuroimaging morphological findings?: A voxel-based meta-analysis: Dissociability of autism, OCD, schizophrenia. Autism Res 2017; 10: 1079–1095.

15 Chen H, Uddin LQ, Duan X, Zheng J, Long Z, Zhang Y et al. Shared atypical default mode and salience network functional connectivity between autism and schizophrenia: Shared atypical FC in ASD and schizophrenia. Autism Res 2017; 10: 1776–1786.

16 Scott-Van Zeeland AA, Dapretto M, Ghahremani DG, Poldrack RA, Bookheimer SY. Reward processing in autism. Autism Res 2010; 3: 53–67.

17 Kesby JP, Murray GK, Knolle F. Neural Circuitry of Salience and Reward Processing in Psychosis. Biol Psychiatry Glob Open Sci 2021. doi:10.1016/j.bpsgos.2021.12.003.

18 Kinard JL, Mosner MG, Greene RK, Addicott M, Bizzell J, Petty C et al. Neural Mechanisms of Social and Nonsocial Reward Prediction Errors in Adolescents with Autism Spectrum Disorder. Autism Res 2020; 13: 715–728.

19 Cannon J, O’Brien AM, Bungert L, Sinha P. Prediction in Autism Spectrum Disorder: A Systematic Review of Empirical Evidence. Autism Res 2021; 14: 604–630.

20 Montagnese M, Knolle F, Haarsma J, Griffin JD, Richards A, Vertes PE et al. Reinforcement learning as an intermediate phenotype in psychosis? Deficits sensitive to illness stage but not associated with polygenic risk of schizophrenia in the general population. Schizophr Res 2020; 222: 389–396.

21 Ermakova AO, Knolle F, Justicia A, Bullmore ET, Jones PB, Robbins TW et al. Abnormal reward prediction-error signalling in antipsychotic naive individuals with first-episode psychosis or clinical risk for psychosis. Neuropsychopharmacology 2018; 43: 1691–1699.

22 Dinsdale NL, Hurd PL, Wakabayashi A, Elliot M, Crespi BJ. How Are Autism and Schizotypy Related? Evidence from a Non-Clinical Population. PLOS ONE 2013; 8: e63316.

23 Zhou H, Yang H, Gong J, Cheung EFC, Gooding DC, Park S et al. Revisiting the overlap between autistic and schizotypal traits in the non-clinical population using meta-analysis and network analysis. Schizophr Res 2019; 212: 6–14.

24 Larson FV, Wagner AP, Chisholm K, Reniers RLEP, Wood SJ. Adding a Dimension to the Dichotomy: Affective Processes Are Implicated in the Relationship Between Autistic and Schizotypal Traits. Front Psychiatry 2020; 11.

25 McCutcheon RA, Krystal JH, Howes OD. Dopamine and glutamate in schizophrenia: biology, symptoms and treatment. World Psychiatry 2020; 19: 15–33.

26 Uno Y, Coyle JT. Glutamate hypothesis in schizophrenia. Psychiatry Clin Neurosci 2019; 73: 204–215.

27 Kaminski J, Mascarell-Maricic L, Fukuda Y, Katthagen T, Heinz A, Schlagenhauf F. Glutamate in the Dorsolateral Prefrontal Cortex in Patients With Schizophrenia: A Meta-analysis of 1H-Magnetic Resonance Spectroscopy Studies. Biol Psychiatry 2021; 89: 270–277.

28 Godlewska BR, Minichino A, Emir U, Angelescu I, Lennox B, Micunovic M et al. Brain glutamate concentration in men with early psychosis: A magnetic resonance spectroscopy Case–Control study at 7 T. Transl Psychiatry 2021; 11: 1–7.

29 Jauhar S, McCutcheon R, Borgan F, Veronese M, Nour M, Pepper F et al. The relationship between cortical glutamate and striatal dopamine in first-episode psychosis: a cross-sectional multimodal PET and magnetic resonance spectroscopy imaging study. Lancet Psychiatry 2018; 5: 816–823.

30 Sydnor VJ, Roalf DR. A meta-analysis of ultra-high field glutamate, glutamine, GABA and glutathione 1HMRS in psychosis: Implications for studies of psychosis risk. Schizophr Res 2020; 226: 61–69.

31 Kumar J, Liddle EB, Fernandes CC, Palaniyappan L, Hall EL, Robson SE et al. Glutathione and glutamate in schizophrenia: a 7T MRS study. Mol Psychiatry 2020; 25: 873–882.

32 Moghaddam B, Krystal JH. Capturing the angel in ‘angel dust’: twenty years of translational neuroscience studies of NMDA receptor antagonists in animals and humans. Schizophr Bull 2012; 38: 942–949.

33 Haarsma J, Knolle F, Griffin JD, Taverne H, Mada M, Goodyer IM et al. Influence of Prior Beliefs on Perception in Early Psychosis: Effects of Illness Stage and Hierarchical Level of Belief. J Abnorm Psychol 2020; 129: 581–598.

34 Zahid U, Onwordi EC, Hedges EP, Wall MB, Modinos G, Murray RM et al. Neurofunctional correlates of glutamate and GABA imbalance in psychosis: A systematic review. Neurosci Biobehav Rev 2023; 144: 105010.

35 Horder J, Petrinovic MM, Mendez MA, Bruns A, Takumi T, Spooren W et al. Glutamate and GABA in autism spectrum Disorder—a translational magnetic resonance spectroscopy study in man and rodent models. Transl Psychiatry 2018; 8: 1–11.

36 Page LA, Daly E, Schmitz N, Simmons A, Toal F, Deeley Q et al. In vivo 1H-magnetic resonance spectroscopy study of amygdala-hippocampal and parietal regions in autism. Am J Psychiatry 2006; 163: 2189–2192.

37 Sapey-Triomphe L-A, Temmerman J, Puts NAJ, Wagemans J. Prediction learning in adults with autism and its molecular correlates. Mol Autism 2021; 12: 64.

38 He JL, Oeltzschner G, Mikkelsen M, Deronda A, Harris AD, Crocetti D et al. Region-specific elevations of glutamate + glutamine correlate with the sensory symptoms of autism spectrum disorders. Transl Psychiatry 2021; 11: 1–10.

39 Doyle-Thomas KAR, Card D, Soorya LV, Ting Wang A, Fan J, Anagnostou E. Metabolic mapping of deep brain structures and associations with symptomatology in autism spectrum disorders. Res Autism Spectr Disord 2014; 8: 44–51.

40 Joshi G, Biederman J, Wozniak J, Goldin RL, Crowley D, Furtak S et al. Magnetic resonance spectroscopy study of the glutamatergic system in adolescent males with high-functioning autistic disorder: a pilot study at 4T. Eur Arch Psychiatry Clin Neurosci 2013; 263: 379–384.

41 Brown MS, Singel D, Hepburn S, Rojas DC. Increased Glutamate Concentration in the Auditory Cortex of Persons With Autism and First-Degree Relatives: A ^1^ H-MRS Study: Increased glutamate concentration in autism. Autism Res 2013; 6: 1–10.

42 Tebartz van Elst L, Maier S, Fangmeier T, Endres D, Mueller GT, Nickel K et al. Disturbed cingulate glutamate metabolism in adults with high-functioning autism spectrum disorder: evidence in support of the excitatory/inhibitory imbalance hypothesis. Mol Psychiatry 2014; 19: 1314–1325.

43 Kolodny T, Schallmo M-P, Gerdts J, Edden RAE, Bernier RA, Murray SO. Concentrations of Cortical GABA and Glutamate in Young Adults With Autism Spectrum Disorder. Autism Res Off J Int Soc Autism Res 2020; 13: 1111–1129.

44 Shirayama Y, Matsumoto K, Osone F, Hara A, Guan S, Hamatani S et al. The Lack of Alterations in Metabolites in the Medial Prefrontal Cortex and Amygdala, but Their Associations with Autistic Traits, Empathy, and Personality Traits in Adults with Autism Spectrum Disorder: A Preliminary Study. J Autism Dev Disord 2022. doi:10.1007/s10803-022-05778-7.

45 Maier S, Düppers AL, Runge K, Dacko M, Lange T, Fangmeier T et al. Increased prefrontal GABA concentrations in adults with autism spectrum disorders. Autism Res 2022; 15: 1222–1236.

46 Li W, Pozzo-Miller L. Dysfunction of the corticostriatal pathway in autism spectrum disorders. J Neurosci Res 2020; 98: 2130–2147.

47 Dandash O, Pantelis C, Fornito A. Dopamine, fronto-striato-thalamic circuits and risk for psychosis. Schizophr Res 2017; 180: 48–57.

48 Sabaroedin K, Razi A, Chopra S, Tran N, Pozaruk A, Chen Z et al. Effective Connectivity of Fronto-Striato-Thalamic Circuitry Across the Psychosis Continuum. Biol Psychiatry 2021; 89: S356.

49 Ford TC, Crewther DP. A Comprehensive Review of the 1H-MRS Metabolite Spectrum in Autism Spectrum Disorder. Front Mol Neurosci 2016; 9. doi:10.3389/fnmol.2016.00014.

50 Shukla DK, Wijtenburg SA, Chen H, Chiappelli JJ, Kochunov P, Hong LE et al. Anterior Cingulate Glutamate and GABA Associations on Functional Connectivity in Schizophrenia. Schizophr Bull 2019; 45: 647–658.

51 Ford TC, Abu-Akel A, Crewther DP. The association of excitation and inhibition signaling with the relative symptom expression of autism and psychosis-proneness: Implications for psychopharmacology. Prog Neuropsychopharmacol Biol Psychiatry 2019; 88: 235–242.

52 Wenneberg C, Glenthøj BY, Hjorthøj C, Buchardt Zingenberg FJ, Glenthøj LB, Rostrup E et al. Cerebral glutamate and GABA levels in high-risk of psychosis states: A focused review and meta-analysis of 1H-MRS studies. Schizophr Res 2020; 215: 38–48.

53 Purcell AE, Jeon OH, Zimmerman AW, Blue ME, Pevsner J. Postmortem brain abnormalities of the glutamate neurotransmitter system in autism. Neurology 2001; 57: 1618–1628.

54 Nelson MT, Seal ML, Pantelis C, Phillips LJ. Evidence of a dimensional relationship between schizotypy and schizophrenia: a systematic review. Neurosci Biobehav Rev 2013; 37: 317–327.

55 Chisholm K, Lin A, Abu-Akel A, Wood SJ. The association between autism and schizophrenia spectrum disorders: A review of eight alternate models of co-occurrence. Neurosci Biobehav Rev 2015; 55: 173–183.

56 Zhou H, Yang H, Gong J, Cheung EFC, Gooding DC, Park S et al. Revisiting the overlap between autistic and schizotypal traits in the non-clinical population using meta-analysis and network analysis. Schizophr Res 2019; 212: 6–14.

57 Ford TC, Nibbs R, Crewther DP. Glutamate/GABA+ ratio is associated with the psychosocial domain of autistic and schizotypal traits. PLOS ONE 2017; 12: e0181961.

58 Klein C, Andresen B, Jahn T. Erfassung der schizotypen Persönlichkeit nach DSM-III-R: Psychometrische Eigenschaften einer autorisierten deutschsprachigen Übersetzung des ‘Schizotypal Personality Questionnaire’ (SPQ) von Raine. [Psychometric assessment of the schizotypal personality according to DSM-III-R criteria: Psychometric properties of an authorized German translation of Raine’s ‘Schizotypal Personality Questionnaire’ (SPQ).]. Diagnostica 1997; 43: 347–369.

59 Raine A. The SPQ: A Scale for the Assessment of Schizotypal Personality Based on DSM-III-R Criteria. Schizophr Bull 1991; 17: 555–564.

60 Wuthrich V, Bates TC. Confirmatory Factor Analysis of the Three-Factor Structure of the Schizotypal Personality Questionnaire and Chapman Schizotypy Scales. J Pers Assess 2006; 87: 292–304.

61 Baron-Cohen S, Wheelwright S, Skinner R, Martin J, Clubley E. The Autism-Spectrum Quotient (AQ): Evidence from Asperger Syndrome/High-Functioning Autism, Malesand Females, Scientists and Mathematicians. J Autism Dev Disord 2001; 31: 5–17.

62 Wilson M. spant: An R package for magnetic resonance spectroscopy analysis. J Open Source Softw 2021; 6: 3646.

63 Landheer K, Swanberg KM, Juchem C. Magnetic resonance Spectrum simulator (MARSS), a novel software package for fast and computationally efficient basis set simulation. NMR Biomed 2021; 34: e4129.

64 Provencher S. LCModel1 & LCMgui User’s Manual. 2021.http://lcmodel.ca/pub/LCModel/manual/manual.pdf.

65 Wilson M. Adaptive baseline fitting for MR spectroscopy analysis. Magn Reson Med 2021; 85: 13–29.

66 Clarke WT, Stagg CJ, Jbabdi S. FSL-MRS: An end-to-end spectroscopy analysis package. Magn Reson Med 2021; 85: 2950–2964.

67 Gasparovic C, Song T, Devier D, Bockholt HJ, Caprihan A, Mullins PG et al. Use of tissue water as a concentration reference for proton spectroscopic imaging. Magn Reson Med 2006; 55: 1219–1226.

68 Geurts JJG, Barkhof F, Castelijns JA, Uitdehaag BMJ, Polman CH, Pouwels PJW. Quantitative 1H-MRS of healthy human cortex, hippocampus, and thalamus: Metabolite concentrations, quantification precision, and reproducibility. J Magn Reson Imaging 2004; 20: 366–371.

69 Akoglu H. User’s guide to correlation coefficients. Turk J Emerg Med 2018; 18: 91–93.

70 Gu Z, Gu L, Eils R, Schlesner M, Brors B. circlize implements and enhances circular visualization in R. Bioinformatics 2014; 30: 2811–2812.

71 Modinos G, McLaughlin A, Egerton A, McMullen K, Kumari V, Barker GJ et al. Corticolimbic hyper-response to emotion and glutamatergic function in people with high schizotypy: a multimodal fMRI-MRS study. Transl Psychiatry 2017; 7: e1083–e1083.

72 Merritt K, Egerton A, Kempton MJ, Taylor MJ, McGuire PK. Nature of Glutamate Alterations in Schizophrenia: A Meta-analysis of Proton Magnetic Resonance Spectroscopy Studies. JAMA Psychiatry 2016; 73: 665–674.

73 Sigvard AK, Bojesen KB, Ambrosen KS, Nielsen MØ, Gjedde A, Tangmose K et al. Dopamine Synthesis Capacity and GABA and Glutamate Levels Separate Antipsychotic-Naïve Patients With First-Episode Psychosis From Healthy Control Subjects in a Multimodal Prediction Model. Biol Psychiatry Glob Open Sci 2022; : S2667174322000659.

74 Reid MA, Salibi N, White DM, Gawne TJ, Denney TS, Lahti AC. 7T Proton Magnetic Resonance Spectroscopy of the Anterior Cingulate Cortex in First-Episode Schizophrenia. Schizophr Bull 2019; 45: 180–189.

75 Marsman A, van den Heuvel MP, Klomp DWJ, Kahn RS, Luijten PR, Hulshoff Pol HE. Glutamate in schizophrenia: a focused review and meta-analysis of ^1^H-MRS studies. Schizophr Bull 2013; 39: 120–129.

76 Maximo JO, Briend F, Armstrong WP, Kraguljac NV, Lahti AC. Salience network glutamate and brain connectivity in medication-naïve first episode patients – A multimodal magnetic resonance spectroscopy and resting state functional connectivity MRI study. NeuroImage Clin 2021; 32: 102845.

77 Modinos G, Egerton A, McLaughlin A, McMullen K, Kumari V, Lythgoe DJ et al. Neuroanatomical changes in people with high schizotypy: Relationship to glutamate levels. Psychol Med 2018; 48: 1880–1889.

78 Demro C, Rowland L, Wijtenburg SA, Waltz J, Gold J, Kline E et al. Glutamatergic metabolites among adolescents at risk for psychosis. Psychiatry Res 2017; 257: 179–185.

79 Egerton A, Brugger S, Raffin M, Barker GJ, Lythgoe DJ, McGuire PK et al. Anterior Cingulate Glutamate Levels Related to Clinical Status Following Treatment in First-Episode Schizophrenia. Neuropsychopharmacology 2012; 37: 2515–2521.

80 Egerton A, Griffiths K, Casetta C, Deakin B, Drake R, Howes OD et al. Anterior cingulate glutamate metabolites as a predictor of antipsychotic response in first episode psychosis: data from the STRATA collaboration. Neuropsychopharmacology 2023; 48: 567–575.

81 Kubota M, Moriguchi S, Takahata K, Nakajima S, Horita N. Treatment effects on neurometabolite levels in schizophrenia: A systematic review and meta-analysis of proton magnetic resonance spectroscopy studies. Schizophr Res 2020; 222: 122–132.

82 Veerman S, Schulte P, de Haan L. The Glutamate Hypothesis: A Pathogenic Pathway from which Pharmacological Interventions have Emerged. Pharmacopsychiatry 2014; 47: 121–130.

83 Overbeek G, Gawne TJ, Reid MA, Kraguljac NV, Lahti AC. A multimodal neuroimaging study investigating resting-state connectivity, glutamate and GABA at 7 T in first-episode psychosis. J Psychiatry Neurosci 2021; 46: E702–E710.

84 Peters H, Riedl V, Manoliu A, Scherr M, Schwerthöffer D, Zimmer C et al. Changes in extra-striatal functional connectivity in patients with schizophrenia in a psychotic episode. Br J Psychiatry 2017; 210: 75–82.

85 Cochrane M, Petch I, Pickering AD. Do measures of schizotypal personality provide non-clinical analogues of schizophrenic symptomatology? Psychiatry Res 2010; 176: 150–154.

86 Kozhuharova P, Diaconescu AO, Allen P. Reduced cortical GABA and glutamate in high schizotypy. Psychopharmacology (Berl) 2021; 238: 2459–2470.

87 Ford TC, Nibbs R, Crewther DP. Increased glutamate/GABA+ ratio in a shared autistic and schizotypal trait phenotype termed Social Disorganisation. NeuroImage Clin 2017; 16: 125–131.

88 Kondo HM, Lin I-F. Excitation-inhibition balance and auditory multistable perception are correlated with autistic traits and schizotypy in a non-clinical population. Sci Rep 2020; 10: 8171.

89 Ford TC, Crewther DP. Factor Analysis Demonstrates a Common Schizoidal Phenotype within Autistic and Schizotypal Tendency: Implications for Neuroscientific Studies. Front Psychiatry 2014; 5.

