## Supplemental files for "Association between increased anterior cingulate glutamate and psychotic-like symptoms, but not autistic traits"

**Running title:** Glutamate, schizotypal and autistic traits

Demler et al.

### Methods

#### Participants

53 healthy participants (28 women, 27 men) aged between 18 to 35 years were recruited through local advertisements and social media. To evaluate eligibility for participating in our study at the Technical University of Munich, all subjects completed a clinical online questionnaire to assess their psychotic-like experiences and autistic traits. Demographic data and medical history were collected during a brief telephone screening to ensure the fulfillment of our inclusion criteria. All participants were German native speakers; right-handed; had no diagnosis of schizophrenia, psychosis or autism, or any neurological disease or injury and were currently not taking any psychoactive medication or had a change in their medication within at least the past six weeks. All participants had no contraindications for MRI scanning. Three subjects had a previous diagnosis of depression, two had previous eating disorders, one was diagnosed as schizoid with adjustment disorder and another subject reported a post-traumatic stress disorder and a narcissistic personality disorder. However, only one participant was taking a stable dose of antipsychotic medication for attention deficit disorder (fluoxetine, atomoxetine). Details of demographic data and symptom scores are shown in Table 1 and Figure S1. The study was approved by the medical research ethics committee of the Technical University of Munich. All subjects gave written informed consent in accordance with the Declaration of Helsinki.

### Schizotypal Personality Questionnaire

The Schizotypal Personality Questionnaire (SPQ) is a widely used 74-item self-report questionnaire to capture psychotic-like experiences according to the DSM-III-R based constructs of schizotypal personality disorder symptoms [1]. It measures nine dimensions of psychotic-like experiences which can be assigned to a three-factor structure consisting of positive-like symptoms (i.e., ideas of reference, magical thinking, unusual perceptual experiences, paranoia), negative-like symptoms (i.e., no closer friends, constricted affect, social anxiety), and disorganized traits (i.e., odd speech, odd behavior; [2]). In its original version, the SPQ is administered in a forced-choice yes/no. Here we applied a German translation [3] of the modified version of the SPQ using a 5-point Likert scale version (strongly disagree=0, disagree=1, neutral=2, agree=3, strongly agree=4) with a maximum of 296 points, which increases the sensitivity of detecting schizotypy traits [4].

### Autism Spectrum Quotient

The Autism Spectrum Quotient (AQ) is an established 50-item self-report questionnaire which is designed to assess five different facets of autistic spectrum traits (social skill, attention switching, attention to detail, communication, imagination) [5]. The AQ is administered in a 4-point Likert scale format (definitely agree, slightly agree, slightly disagree, definitely disagree). For the evaluation, a binary scoring method is used with the presence of autistic traits, either mildly or strongly, generating one point while the opposite is scored zero, leading to a maximum score of 50. The items are counterbalanced so that half of the items are worded to produce a disagree response and the other half to produce an agree response in a high scoring individual. In the present study, we applied a German translation of the AQ scale.

### Results

#### $^1$H-MRS tCr and tNAA levels and spectral quality

For our control analyses, we measured the concentration of total creatine (tCr; creatine + phosphocreatine) and total (tNAA; N-acetylaspartate + N-acetyl-aspartylglutamate) in our five voxels of interest, the ACC, the left/right putamen, and the left/right DLPFC. Results are presented in Table S1.

#### Association between positive-like symptoms and tCr or tNAA concentration in the anterior cingulate cortex

We first fitted a complete multiple linear regression model to test if tCr or tNAA concentrations in our five voxels of interest predicted positive-like symptoms. Contrary to glutamate we did not find any significant predictions (see Table S2).

#### Correlation between grey matter volume and glutamate concentration across different regions

To investigate the relationship between the grey matter volume and glutamate concentration in the different regions, we calculated Spearman's rank correlation coefficients as the assumptions of normality were not met. Results were visualized using a scatter plot (see Figure S1). The scatter plots and correlation analyses were performed using ggpubr version 0.5.0 (https://CRAN.R-project.org/package=ggpubr).

We did not find a correlation between the grey matter volume and the glutamate concentration in the ACC (see Figure S2), indicating that higher glutamate in this region is not due to increased grey matter volume.

### References


1 Raine A. The SPQ: A Scale for the Assessment of Schizotypal Personality Based on DSM-III-R Criteria. *Schizophr Bull* 1991; **17**: 555–564.

2 Wuthrich V, Bates TC. Confirmatory Factor Analysis of the Three-Factor Structure of the Schizotypal Personality Questionnaire and Chapman Schizotypy Scales. *J Pers Assess* 2006; **87**: 292–304.

3 Klein C, Andresen B, Jahn T. Erfassung der schizotypen Persönlichkeit nach DSM-III-R: Psychometrische Eigenschaften einer autorisierten deutschsprachigen Übersetzung des 'Schizotypal Personality Questionnaire' (SPQ) von Raine. [Psychometric assessment of the schizotypal personality according to DSM-III-R criteria: Psychometric properties of an authorized German translation of Raine's 'Schizotypal Personality Questionnaire' (SPQ).]. *Diagnostica* 1997; **43**: 347–369.

4 Wuthrich V, Bates TC. Reliability and validity of two Likert versions of the Schizotypal Personality Questionnaire (SPQ). *Personal Individ Differ* 2005; **38**: 1543–1548.

5 Baron-Cohen S, Wheelwright S, Skinner R, Martin J, Clubley E. The Autism-Spectrum Quotient (AQ): Evidence from Asperger Syndrome/High-Functioning Autism, Malesand Females, Scientists and Mathematicians. *J Autism Dev Disord* 2001; **31**: 5–17.

### Tables

*Table S1: [1]H-MRS quality parameters and metabolite levels by region*

| Region | ACC | PUT R | PUT L | DLPFC R | DLPFC L |
|---|---|---|---|---|---|
| n | 53 | 53 | 53 | 53 | 53 |
| tNAA (SD), mMol/kg | 16.52 (1.14) | 14.45 (2.06) | 14.25 (1.21) | 15.25 (0.92) | 15.76 (1.35) |
| CRLB (SD), % | 0.81 (0.12) | 1.31 (1.18) | 1.29 (0.23) | 0.61 (0.84) | 0.61 (0.14) |
| tCr (SD), mMol/kg | 14.06 (0.97) | 12.14 (0.69) | 11.53 (0.82) | 10.83 (0.81) | 11.11 (0.91) |
| CRLB (SD), % | 0.49 (0.07) | 0.88 (0.18) | 0.91 (0.21) | 0.45 (0.07) | 0.45 (0.09) |

*Note: Values are mean (SD)*

*tCr, total-creatine (Cr, creatine + PCr, phosphocreatine); tNAA, total-N-acetylaspartate (NAA, N-acetylaspartate + NAAG, N-acetyl-aspartylglutamate); ACC, anterior cingulate cortex; PUT R, right putamen; PUT L, left putamen; DLPFC R, right dorsolateral prefrontal cortex; DLPFC L, left dorsolateral prefrontal cortex; Glu, Glutamate, CRLB, Cramer-Rao Lower Bound; SD, standard deviation*

*Table S2: Results of the regression analysis for each metabolite.*

| Metabolite | Model | Statistics | Predictor - ACC |
|---|---|---|---|
| tCr (Cr+PCr) | Positive-like symptoms ~ PUT R + PUT L + DLPFC R + DLPFC L + ACC | F(47, 5)= 0.81, p=0.551, r2=-0.02 | ACC: β=3.81 (3.88), t=0.98, p=0.331 |
| tNAA (NAA+NAAG) | Positive-like symptoms ~ PUT R + PUT L + DLPFC R + DLPFC L + ACC | F(47, 5)=1.52, p=0.203, r2=-0.05 | ACC: β=4.14 (2.95), t=1.41, p= 0.167 |

*Note: tCr, total-creatine; Cr, creatine; PCr, phosphocreatine; tNAA, total-N-acetylaspartate; NAA, N-acetylaspartate; NAAG, N-acetyl-aspartylglutamate; ACC, anterior cingulate cortex, DLPFC L, left dorsolateral prefrontal cortex; DLPFC R, right dorsolateral prefrontal cortex; PUT L, left putamen; PUT R, right putamen*

### Figures

*Figure S1: Distribution of the clinical scores*

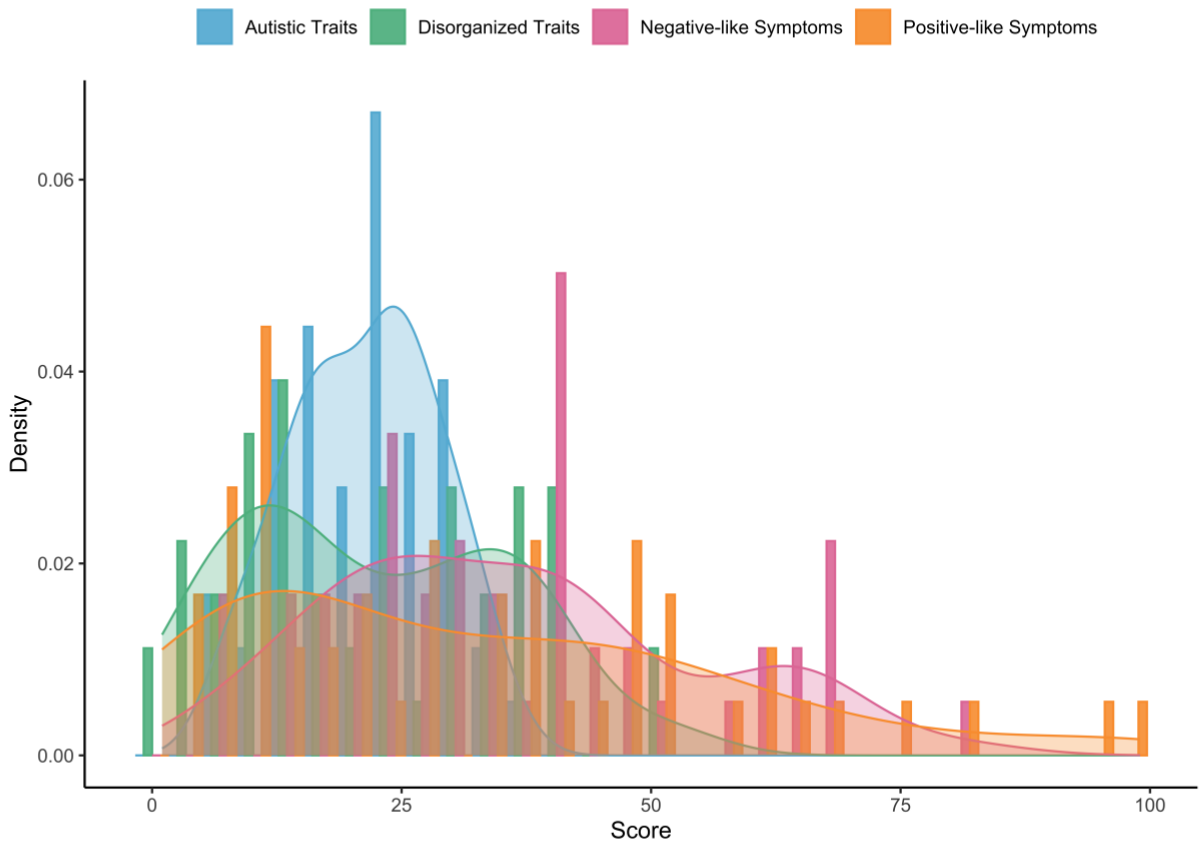


*Note: The histogram shows the distribution of each trait score marked by the different colours. The subscores are structured according to their respective associated traits (i.e., Autistic Traits for AQ; Negative-like Symptoms, Posititive-like Symptoms, and Disorganized traits for SPQ).*

*Figure S2: MRS-data Processing and Analysis workflow*

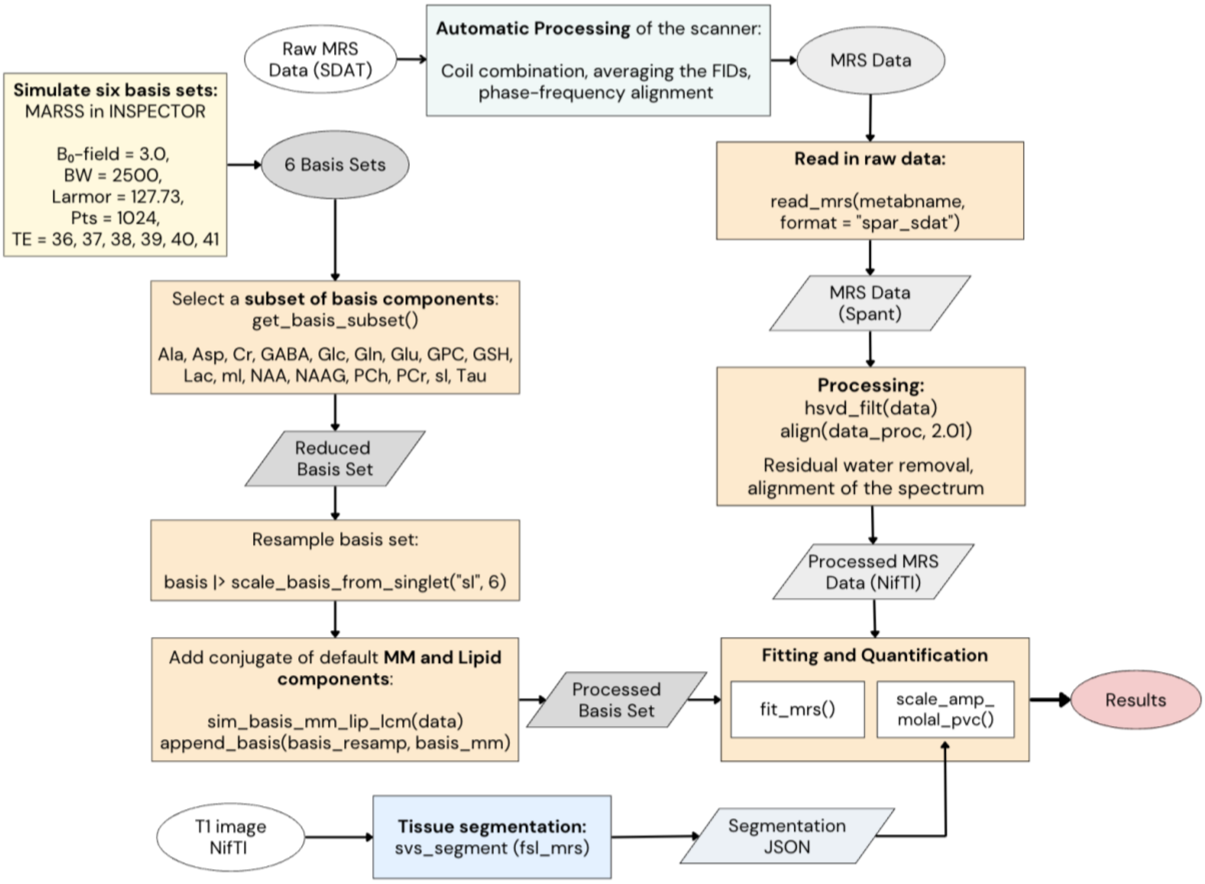


*Note: Workflow of our MRS-data processing, Colors indicate the different used tools; yellow for MARSS in INSPECTOR, orange for spant and blue for fsl mrs;*

*BW, bandwidth; TE, time of echo; MM, makromolecules; Ala, alanine; Asp, aspartate; Cr, creatine; GABA; Glc, glucose; Glu, glutamate; Gln, glutamine; GSH, glutathione; GPC, glycerophosphocholine; Lac, lactate; mI, myoinosito; NAA, N-acetylaspartate; NAAG, N-acetyl-aspartylglutamate; PCh, phosphocholine; PCr, phosphocreatine; sI, scyllo-inositol; Tau, taurine*

*Figure S3: Correlation between grey matter and glutamate grouped by brain regions*

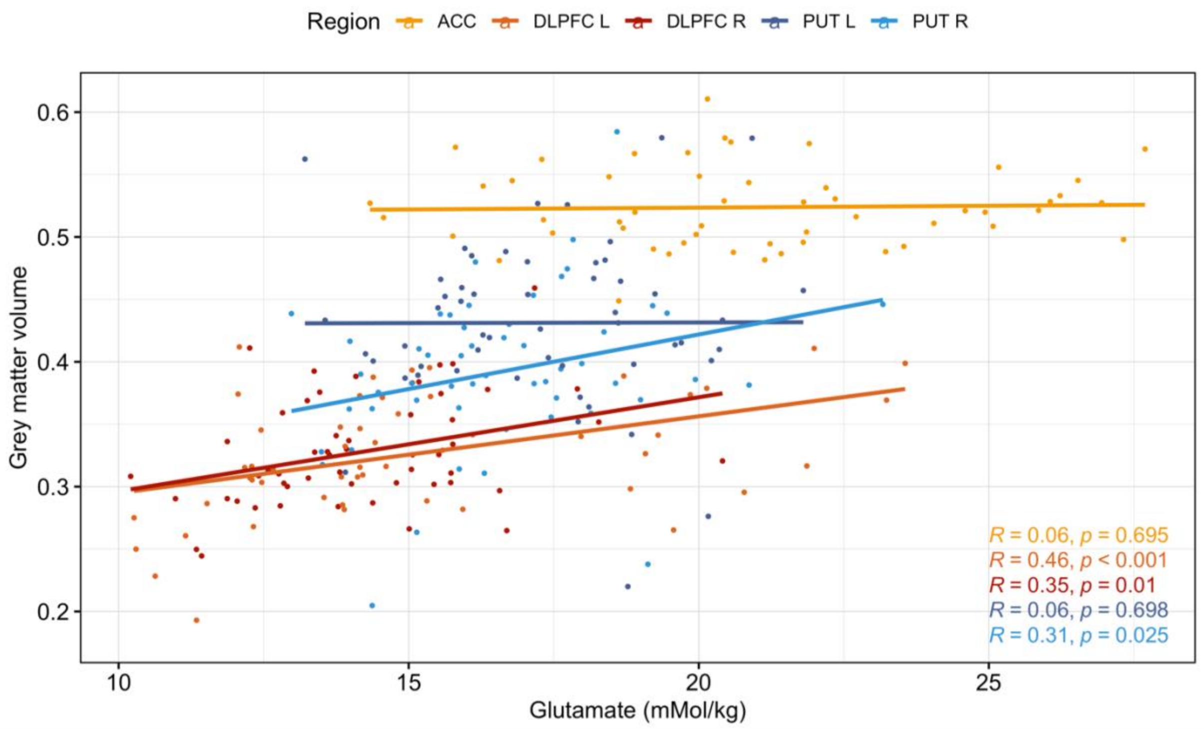


*Note: Scatterplot with correlation coefficients for n=53, whereas the colors indicate the different voxel locations; ACC, anterior cingulate cortex, DLPFC L, left dorsolateral prefrontal cortex; DLPFC R, right dorsolateral prefrontal cortex; PUT L, left putamen; PUT R, right putamen*
